# RSV and Rhinovirus asymptomatic upper airway infection increases pneumococcal carriage acquisition rates and density in adults whereas nasal inflammation is associated with bacterial shedding

**DOI:** 10.1101/2024.02.08.24302534

**Authors:** Elena Mitsi, Elissavet Nikolaou, Andre Goncalves, Annie Blizard, Helen Hill, Madlen Farrar, Angela Hyder-Wright, Ashleigh Howard, Filora Elterish, Carla Solorzano, Ryan Robinson, Jesus Reine, Andrea M. Collins, Stephen B. Gordon, Jeffrey N. Weiser, Debby Bogaert, Daniela M. Ferreira

## Abstract

Nasopharyngeal pneumococcal colonisation is a necessary step for disease development and the primary reservoir of bacterial spread and transmission. Most epidemiological studies report the impact of co-infection with respiratory viruses upon disease rates and outcome, but their effect on pneumococcal carriage acquisition and bacterial load is scarcely described. Here, we used controlled human infection with pneumococcus to assess whether certain respiratory viruses alter susceptibility to pneumococcal colonisation and bacterial density. A total of 581 healthy adults were screened for presence of upper respiratory tract viral infection before intranasal pneumococcal challenge. We showed that RSV and Rhinovirus asymptomatic infection conferred a substantial increase in carriage rates (88% and 66% of colonised individuals in Rhinovirus and RSV infected groups, respectively, vs 49% in the virus negative group). The risk ratio of pneumococcal colonization in RSV infected group was 5.3 (95% CI: 0.90 – 30.61, p = 0.034) and 2.03 (95% CI: 1.09-3.80, p= 0.035) in rhinovirus infected group. Pneumococcal density was overall greater in virus positive subjects, although RSV infection alone had a major impact on pneumococcal density up to 9 days post challenge, with a substantial 2- log increase at D2 compared to virus negative group (median, SEM: 3.76 ± 0.65 vs 1.79 ± 0.09) (p= 0.02). We also studied rates and kinetics of bacterial shedding through the nose and oral route in a subset of challenged individuals. Nasal bacterial shedding was twice more frequent than cough-induced shedding (64% vs 32%, respectively). High levels of bacterial colonisation density and nasal inflammation was strongly correlated with increased odds of nasal shedding, as opposed to cough-shedding.

Healthy adults can act as reservoir of transmission for pneumococcus, particularly when colonised with high density and have local inflammation, two key characteristics of pneumococcal colonisation in paediatrics and/or viral co-infection. Hence, vaccines targeting these respiratory pathogens have the dual potential of reducing transmission and disease burden due to their indirect benefit to off target pathogens.

## INTRODUCTION

*Streptococcus pneumoniae* (the pneumococcus) is a leading opportunistic pathogen of the human respiratory tract. Nasopharyngeal pneumococcal colonisation is a common event, with 40-90% prevalence in young children (<5 years old) and 10-20% in adults in epidemiological studies [1–4]. Although colonisation of the nasopharynx is generally considered asymptomatic (the carrier state), and young children experience multiple colonisation episodes, mostly without incident, under certain circumstances pneumococci can invade sites and spread within the host, resulting in disease development[5–7]. This paradox suggests that other factors beyond nasopharyngeal carriage play an important role in the pathogenesis of pneumococcal disease [8, 9].

Viral infections, mainly respiratory syncytial virus (RSV) and influenza, seem to increase substantially the risk of secondary pneumococcal infection, including invasive pneumococcal disease (IPD) [10, 11]. Epidemiological, clinical and experimental evidence suggest that these viruses increase bacterial virulence and worsening disease outcome [12, 13], implying that non-invasive pneumococcal serotypes possibly require enhancement by respiratory viruses to cause secondary bacterial infection.

Colonisation is also the primary source and reservoir of pneumococcal transmission [6]. It is postulated that a colonised person can shed bacteria via nasal secretion and mucosalivary fluid in the form of droplets [14]. Colonisation and transmission are most common among young children, especially in crowded settings, such as day care centres [15], or in association with episodes of viral upper respiratory tract infection [16–18]. In animal models, higher density of colonising bacteria and increased nasal secretions triggered by influenza, leads to increased bacterial shedding [19]. In humans, influenza infection induced by Live attenuated influenza vaccine can trigger acute nasal inflammation [20] and lead to higher density of colonising pneumococci [20, 21]. In children, viral rhinitis has also been associated with elevated pneumococcal colonising density in children [22].

Here we studied the relationship between certain respiratory viruses and predisposition to pneumococcal colonisation using a controlled human infection model with pneumococcus in adults. As the onset of colonisation is known, it ascertains pathogen order and causality on bacterial density and host responses.

In addition, we described the kinetics of bacterial shedding during early and later stages of colonisation and provided first evidence that adults can also act as reservoir of bacterial spread while they are colonised with *S. pneumoniae*.

## METHODS

### Study design and sample collection

A total of 581 healthy adults (aged 18-55 years), who participated in experimental human pneumococcal challenge studies in the Liverpool School of Tropical Medicine, UK, between November 2011 and October 2019, were challenged intranasally with live pneumococci serotype 6B, as previously described [23]. All studies were approved by North-West National Health Service Research Ethics Committee and participants gave informed consent. All participants were non-smoking adults who had no close contact with at-risk individuals, such as young children (under the age of 5 years) and the elderly (over 65 years with comorbidities). The median age of the subjects was 21 (IQR:19-22) and 60% were female (351/581) (Table 1).

**Table 1:** Characteristics of study participants.

|  | Uninfected<br>(n=482) | Asymptomatic URTI<br>(n=99) |  |  |  |  |
| --- | --- | --- | --- | --- | --- | --- |
| **Age in years** |  |  |  |  |  |  |
| Median and IQR | 21 (19-22) | 21 (19-22) |  |  |  |  |
| **Gender** |  |  |  |  |  |  |
| Male (%) | 39%<br>(188/482) | 42% (42/99) |  |  |  |  |
| **Pneumococcal Carriage (%)** |  |  |  |  |  |  |
| Experimental carriage acquisition | 49%<br>(235/482) | 57% (56/99) |  |  |  |  |
| **Viral Infection** | N/A | RH<br>(n=44) | HCoVs<br>(N=25) | RSV<br>(n=8) | HPIVs<br>(n=7) | Others<br>(n=15) |
| Viral strains detected |  |  | NL63, HKU1,<br>OC43 & 229E | RSV-A &<br>RSV-B |  | Adenovirus =4<br>Bocavirus= 4<br>Enterovirus=3<br>HMPV= 2<br>Influenza A/B=2 |
| Male (%) |  | 17 (39%) | 11 (44%) | 4 (50%) | 4 (57%) | 6 (40%) |
| Experimental Carriers (%) |  | 29 (66%) | 11 (44%) | 7 (88%) | 5 (71%) | 4 (27%) |
| Odds Ratio (95% CI) |  |  |  |  |  |  |

Nasal wash samples were collected before the intranasal inoculation (screening visit) and at several time points post inoculation, whereas oropharyngeal swabs were collected only at baseline. For a subset of participants, nasal lining fluid (using Nasosorption^TM^ filters) and nasal cells (using Rhino-Pro curettes) were collected at baseline and on days 2, 7 and 16 after challenge. Also, shedding samples, such as cough plates and nose-to-hand (NtH) swabs were collected for a subset of participants on days 2, 6, 16 and 22 after inoculation with pneumococcus serotype 6B.

### Pneumococcal colonisation and bacterial shedding

Pneumococcal detection was performed in NW samples by classic microbiology methods, as previously described[23, 24]. Participants in whom experimental pneumococci were detected in NW samples at any visit post inoculation were defined as experimentally colonised.

To study bacterial shedding during experimental colonisation, we implemented sampling methodologies, such as cough plate method and nose-to-hand swabbing, which allowed us to evaluate longitudinally bacterial shedding from the oral and nasal route, respectively (Supplementary Fig.1). Post inoculation (at D2, D6, D14, D16 and D22) participants were asked to a) cough twice on a blood agar plate enriched with 4 μg/ml gentamicin (Sigma, UK) and b) rube their nose using the anatomical snuff box of their hands. Then, the snuff box area was swabbed using a sterile cotton swab, following storage in STGG until sample processing. Both types of shedding samples were collected before nasal wash or nasosorption samples collection to avoid artificial induction and pneumococcal shedding was determined by classical microbiology methods in real time. The cough plates were incubated immediately at 37°C with 5% CO_2_ for 24 hours. The NtH swabs were streaked out on a blood agar plate and an additional 20ul of the collected sample in STGG was plated on a separate plate enriched with gentamicin (Sigma, UK) and incubated overnight at 37°C with 5% CO_2_. Following incubation, all the plates were inspected for presence of pneumococci and pneumococcal phenotype identification. Serotype was confirmed by latex agglutination. NtH samples were also tested for presence of pneumococcal DNA by a sequential single-plex quantitative PCR (qPCR) using primers for *lytA* gene and cpr gene, specific for serotypes 6A/B, as previously published [25–27].

### DNA/RNA extraction and viral qPCR

Nucleic acids for viral qPCR were extracted from one aliquot of 250 µL oropharyngeal swab and/or 150ul of a paired nasal wash sample collected at baseline (5 days before inoculation) using the Purelink^TM^ Viral RNA/DNA Mini Kit (Life Technologies Corporation, Carlsbad, CA, USA) according to the manufacturer’s instructions. We tested for a broad panel of respiratory viruses using primers, probes and PCR assay conditions specific for adenoviruses, parainfluenza virus 1–4, human bocavirus, human coronavirus OC43, NL63 and 229E, respiratory syncytial virus (A and B), human metapneumovirus, human rhinoviruses, enteroviruses and human influenza virus A and B, as previously described[28].

### Cytokines analysis in nasal lining fluid

Human nasal cytokines and chemokines were quantified in nasal wash supernatant using a customised cytokine storm Olink Flex 21 target panel (TNFa, IL1a, IL1b, IL6, IL8, IL2, IFNγ, IFNα2, IFNβ1, IFNλ1, IL12, granzyme A, IP-10, MIG, MCP-1, MIP-1a, MIP-1b, GM-CSF, IL33, IL4, IL10) from Olink (Uppsala, Sweden) or a 30-Plex Human Cytokine Magnetic Luminex Kit (Thermo Fisher Scientific), as previously described [20]. On the Olink platform the target protein bound to the double oligonucleotide-labelled antibody probe with high specificity, and then a microfluidic real-time PCR amplification of the oligonucleotide sequence was used to quantitatively detect the resulting DNA sequence. The resulting threshold cycle (Ct)-data were processed for quality control and normalized, using internal and external controls. On the Luminex platform, NW samples were analysed on a LX200 (Bio-Rad) with xPonent3.1 software (Luminex Corp) following the manufacturer’s instructions. All samples were run in duplicate, and standards were run on all plates. Calibration and verification beads were run prior to all runs. All cytokines’ data (both Olink and Luminex panel) were expressed in pg/ml.

### Nasal cell collection and immunophenotyping

Nasal microbiopsies were collecting by nasal curettage (Rhino-pro®, Arlington Scientific), as described previously[29]. Two curette samples were collected into PBS + 5mM EDTA and 0.5% heat inactivated FBS for immunophenotyping and an additional two curette samples were collected into RLT (Qiagen) and stored at −80 for transcriptome analysis. In the microbiopses, nasal epithelial cells and immune cells were stained with LIVE/DEAD® Fixable Violet Dead Cell Stain (Thermofisher), followed by a monoclonal antibody cocktail (Supplementary Table 1). Samples were acquired on a LSR II cytometer (BD Biosciences) and analysed using Flowjo software version 10 (Treestar). Compensation matrices were set using compensation beads (BD Biosciences and Thermofisher). Samples with less than 500 CD45+ leukocytes or 250 EpCam+ epithelial cells were excluded from analysis. To adjust for the variability of cells collected within and between study participants, absolute cell counts of the target populations were normalized with the absolute number of epithelial cells. The gating strategy is shown in Supplementary Fig. 1.

### Nasal Cell RNA Sequencing and Bioinformatic Analysis

#### RNA sequencing

Nasal cells were collected in RLT (QIAGEN) and stored at −80°C until extraction. RNA extraction (RNeasy; QIAGEN), sample integrity assessment (Bioanalyzer; Agilent 2100), library preparation and RNA sequencing (BGISEQ-500RS) were performed at the Beijing Genome Institute. RNA mapping and annotation: Quality control of raw sequencing data was conducted using the FastQC tool. The human reference genome assembly GRCh38 was mapped using STAR 2.5.0a [30]. Read counts from binary alignment map (BAM) files were obtained using featureCounts [31] utilizing a general transfer format (GTF) gene annotation from the Ensembl database [32]. The R/Bioconductor package DESeq2 (version 1.34.0) was used for identifying differentially expressed genes, excluding features absent in more than 75% of samples[33].

#### Functional analysis

Gene Set Enrichment Analysis (GSEA) was conducted to identify significant pathways in the ranked transcriptome (log2 fold-change multiplied by −log10 p-values), independent of the differential expression cut-off [34], using the R package fgsea (version 1.20.0) [35]. Pathways with a p-value < 0.05 and an enrichment of at least five genes were considered statistically significant. The Reactome database (release 79) and Pathway Browser (version 3.750) were used for this analysis [36]. Heatmap plots were generated using the R package pheatmap (version 1.0.12).

### Statistical analysis and Correlation plots

Participant characteristics were summarised as n, median (interquartile range) or frequency (percentage). Chi-squared test and Fisher’s exact test were conducted to identify any significant changes in categorical variables. Non-parametric Wilcoxon paired tests and Mann-Whitney tests were conducted to compare quantitative data within the same group or between two groups, respectively. In addition, Kruskal–Wallis rank-sum test with Dunn’s correction were performed to compare quantitative data amongst groups (three groups comparison). Tests were two-sided with an α level of 0.05. For correlations, two-tailed Pearson’s or Spearman’s *r* test was used. To explore the association between time after infection and vaccination, we employed a linear regression model. Data were analysed in R software version 4.0.3 (R Foundation for Statistical Computing, Vienna, Austria), using rstatix package (version 4.2.3) or in Graphpad Prism version 9.0.

Correlograms plotting the Spearman rank correlation coefficient (r), between all parameter pairs were created with the corrplot package (v0.84) running under R (v3.6.1) in R studio (1.1.456). Clustering of parameters was performed using the ‘hclust’ option of corrMatOrder. Spearman rank two-tailed P values were calculated using corr.test (psych v1.8.12) and graphed (ggplot2 v3.1.1) based on ∗ p < 0.05, ∗∗p < 0.01, ∗∗∗ p < 0.001.

## RESULTS

### Asymptomatic viral infection of the upper airways with RSV and rhinovirus predisposes to pneumococcal carriage acquisition

Cross-sectional analysis of naso/oropharyngeal samples collected from healthy adults in the UK revealed that 17% (99/581) of the individuals were asymptomatically infected with a respiratory virus before pneumococcal challenge (Figure 1A). These asymptomatic URTIs were more frequent during autumn and winter months (October – January) (Suppl. Figure 3A). Only infections caused by single virus were detected, with rhinovirus (RH) being the most frequent (7.6%), followed by seasonal coronaviruses (hCoV, 4%), whereas RSV-A/RSV-B, parainfluenza viruses (PIVs) and adenovirus were less frequently detected (Figure 1A and Table 1), possibly as they are frequently symptomatic and therefore participants would have been excluded prior to enrolment.

**Figure 1.**
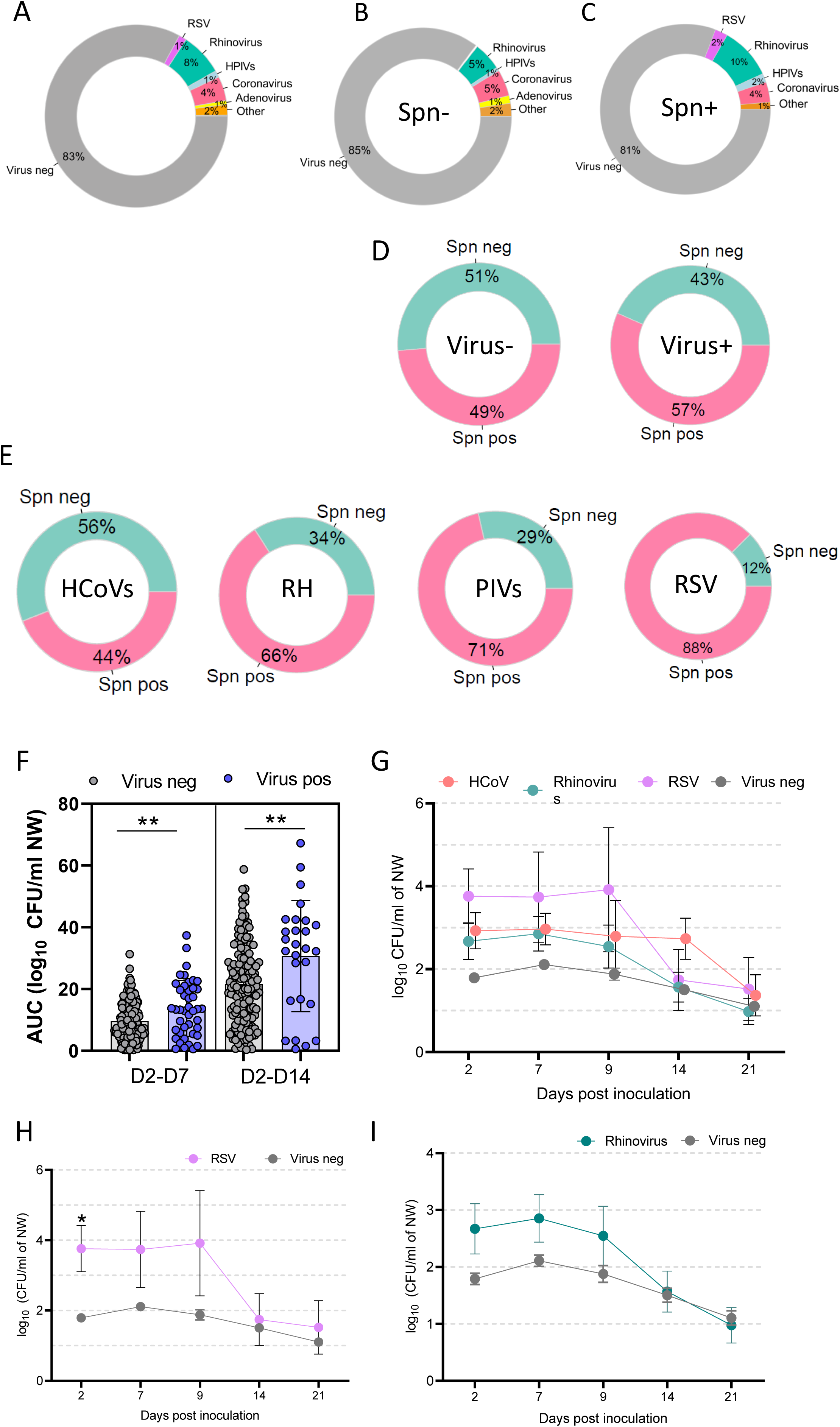
Presence of asymptomatic upper airway viral infection influences pneumococcal carriage acquisition and bacterial density. . **A)** Percentages of respiratory viruses detected in the upper airways of 581 healthy adults at baseline (prior to pneumococcal challenge). Differential distribution of respiratory viruses detected in those who did not become colonised (n=290, Spn- group) and those who became colonised (n=291, Spn+ group). **B)** Percentages of pneumococcal colonisation achieved after experimental challenge in the virus negative (n=482) and virus-positive group (n=99). **C)** Percentages of experimental pneumococcal colonisation achieved after challenge in those with primary asymptomatic seasonal coronavirus (HCoV) infection (11 Spn+ out of 25, p=0.55), rhinovirus (RH) infection (29 Spn+ out of 44, p=0.035), parainfluenza virus (HPIVs) infection (5 Spn+ out of 7, p=0.25) and respiratory syncytial virus (RSV) infection (7 Spn+ out of 8, p=0.034). **D-G)** Density dynamics after pneumococcal inoculation were calculated from classical microbiology (log10[cfu/ml +1]). **D)** Area under the curve (AUC) of density per time interval from day2 to day14 post inoculation with Spn6B in virus negative (n=208) and virus positive (n=43) individuals. **E-G)** Mean ± SEM of pneumococcal density in colonised individuals for each time point after inoculation in those who had primary asymptomatic viral infection with seasonal coronavirus (HCoV, red) or rhinovirus (RH, green) or RSV (purple) or no viral infection (Virus neg, grey). Statistical analysis was determined by Fisher’s exact test to compare percentages (D-E) or Mann-Whitney test (F,H & I) or Kruskal-Wallis test following correction for multiple comparisons (G). p < 0.05, **p < 0.01.

Experimentally induced pneumococcal colonisation was detected in 291 out of the 581 participants (50%), as assessed by longitudinal analysis of the nasal wash samples post challenge and was more common among female participants (56% female vs 41% male, p=0.009), as reported previously [37]. Analysis of the asymptomatic URT cases based on carriage outcome (Spn-colonised and non-colonised group) showed a differential distribution of respiratory viruses between the two groups. RH, RSV and PIV were more apparent in those that become colonised after challenge (Figure 1C) compared to those who remained non-colonised (Figure 1B). In presence of any respiratory virus in the naso/oropharynx prior to challenge, pneumococcal carriage increased from 49% (235 out of 482 virus-negative individuals) to 57% (56 out of 99 virus-positive individuals) (Figure 1D). Assessment of the effect of individual viral species on carriage acquisition revealed that URTI with certain viruses, such as rhinovirus and RSV, likely PIVs but not HCoVs predispose individuals to pneumococcal colonisation (Figure 1E). RH-, RSV- and PIV-infected groups reached colonisation rates of 66% (29/44, p=0.035), 88% (7/8, p=0.035) and 71% ((5/7, p=0.26), respectively, compared to 49% observed in the virus negative group. The risk ratio of pneumococcal colonization was 2.03 (95% CI: 1.09-3.80, p= 0.035) in the RH infected group, 5.3 (95% CI: 0.90 – 30.61, p = 0.034) in the RSV infected group and 2.63 (95% CI: 0.56 – 13.3, p=0.26) in the PIV infected group (Table 2).

**Table 2:** 6B Pneumococcal colonisation status assessed according to viral infection and type of virus.

| Virus | N of colonised/ Total N (%) |  | Odds Ratio (95% CI) | P value |
| --- | --- | --- | --- | --- |
|  | Virus infected | Non-infected |  |  |
| All viruses | 56/99 (57%) | 235/428 (49%) | 1.37 (0.88 – 2.11) | 0.185 |
| RH | 29/44 (66%) | 235/428 (49%) | 2.03 (1.09 – 3.80) | 0.035 |
| HCoV | 11/25 (44%) | 235/428 (49%) | 0.83 (0.37 – 1.78) | 0.55 |
| RSV | 7/8 (88%) | 235/428 (49%) | 5.26 (0.90 – 30.6) | 0.034 |
| PIV | 5/7 (71%) | 235/428 (49%) | 2.63 (0.56 – 13.3) | 0.26 |

We also investigated whether URT viral infection influences the pneumococcal colonising density. The area under the curve (AUC) of pneumococcal colonising density was significantly higher in the virus positive compared to the virus negative group for the Day 2 to Day 7 [median, IQR: 13.38 (5.82-20.21) vs 8.47 (5.2-14.15), p=0.007) and Day2 to Day14 interval post challenge [33.12 (16.25-42.16) vs 20.40 (11.01-30.11), p=0.008] (Figure 1F). Comparison of colonisation density kinetics up to D21 post challenge between virus negative individuals and those infected with HCoV, Rhinovirus or RSV suggested that the presence of any respiratory virus can have a transient effect towards increased bacterial density (Figure 2G-I). Amongst different viruses, presence of RSV in the nasopharynx had the most striking effect, as increased pneumococcal density by 2-log at D2 compared to virus negative group (median, SEM: 3.76 ± 0.65 vs 1.79 ± 0.09) (p= 0.02) an observation that continued up to D9 post challenge (Figure 2H). We also observed that seasonality affected colonising density in the viral infected group. Increased prevalence of viral URTI in Autumn and Winter months was followed by an increased in pneumococcal density during the same period of the year (Suppl. Figure 1B).

**Figure 2.**
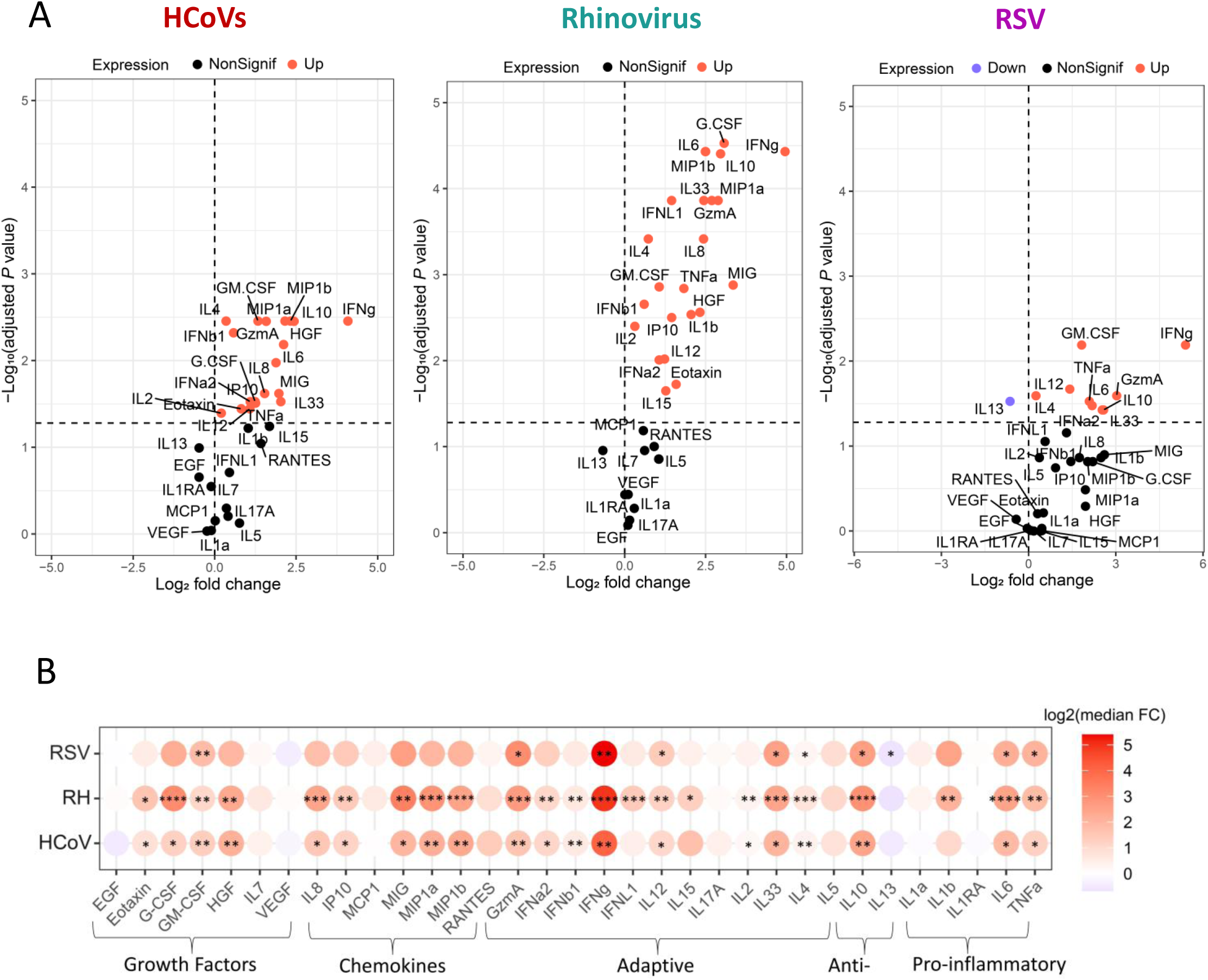
Inflammatory profile of the human upper airway during asymptomatic URTI. **A)** Volcano plots and **B)** heatmap showing the median log_2_ fold change (FC) in the levels of 33 cytokines in the nasal wash collected at baseline from individuals with HCoV (n = 20), rhinovirus (n = 25) and RSV infection (n = 8) versus non-viral infected group (n = 20). On volcano plots the horizontal dotted line represents the cutoff of significance (adjusted p= 0.05, after False Discovery Rate correction of the p value), whereas the vertical dotted line represents a cutoff point for determining whether the levels of cytokines were higher (right, red) or lower (left, blue) compared with those of non-viral infected group. Statistical comparisons were applied between each study group and the virus negative group using the Mann-Whitney U test, followed by Benjamini-Hochberg correction for multiple testing. p < 0.05, **p < 0.01, ***p < 0.001. Nonsignif, nonsignificant; Down, downregulation; Up, upregulation.

### Distinct nasal inflammatory profile in those with asymptomatic URT infection

Levels of nasal inflammation induced by viral infection were assessed by measuring 33 cytokines in the nasal wash samples collected before exposure to pneumococcus (screening). Individuals were divided in three groups based on the type of viral infection (HCoV, RH, RSV) and compared to the non-infected group (virus negative group) (Figure 2). Infection with RH, even in absence of symptoms, elicited an acute nasal inflammatory response, with strong induction of pro-inflammatory mediators (TNFa, IL6 and IL1b), monocyte recruiting chemokines (MIP-1a, MIP-1b) and substantial increase in several antiviral mediators, such as type I, II and II interferons, granzyme A, IP10 and MIG. Asymptomatic infection with seasonal coronaviruses (apha- or beta-coronavirus) induced a similar type of nasal inflammation compared to RH infection, but overall weaker. RSV had a relatively mild inflammatory profile, the least pro-inflammatory amongst the three types of URTI, which was characterized by absence of chemokine induction and a relatively weak antiviral response. Despite of differences introduced due to virus type and infection time, 9 cytokines (GM-CSF, Granzyme A, IFN-g, IL-2, IL33, IL-4, IL10, IL-6 and TNF-a) were identified as commonly induced in all asymptomatic URTIs. Inflammatory responses per group were also stratified and analysed based on the pneumococcal carriage outcome after experimental challenge to investigate whether certain inflammatory mediators are associated with susceptibility to Spn carriage acquisition. In the studied cohort, there was no differentially expressed cytokines between those that would become colonised and those aborted colonisation in each viral group separately or in the combined viral infected groups (data not shown).

### Route of bacterial shedding and association with colonising density

In a subset (n=169) of participants (all individuals enrolled into two consecutive studies from 2018 to 2019) we investigated bacterial shedding through the mouth and nose collecting cough and nose-to-hand samples (Suppl. Figure 1), irrespective of the carriage outcome. Bacterial shedding by any route required successful colonisation after the bacterial challenge at D0. None of the non-colonised individuals (n=48) had a positive shedding sample. Amongst the Spn-colonised (n=121), we identified 47 Spn-shedders, who shed bacterial via the nose and/or mouth at any time point post challenge and 74 non-shedders. In the Spn-shedders group, 64% (30/47) expired bacterial via the nose (nose-shedders), 32% (15/47) via the mouth while coughing (cough-shedders) and only 4% (2/47) via both routes simultaneously (Figure 3A). Bacterial colonising density was consistently higher in the nose-shedders compared to non-shedders group (Figure 3B). Further analysis of bacterial shedding in association with bacterial colonising density revealed a differential profile for cough- and nose- shedders. A gradual increase in the proportion of nose-shedders was observed when bacterial density increased, reaching 80 and 80% in the highest density increments (10^4^-10^5^ and 10^5^-10^6^ CFU/ml of NW, respectively) (Figure 3C and Table 3). In addition, the number of expired bacteria via the nose (shedding density) at any time point was strongly correlated with the nasal bacterial density (Figure 3D) (R=0.52, p=0.0004). Contrarily, cough-shedding was not influenced by fluctuations in nasal bacterial density, as the proportion of cough-shedders remained relatively low in all density increments (Figure 3C and Table 3). This differential pattern was also observed in relation to time post inoculation. Nose-shedders tended to shed bacteria predominately at D2 (72%, 23/32) and D6 (60%, 19/32) post inoculation; time points with highest mean densities (Figure 3F), whereas shedding via cough was occurred evenly across time points (Figure 3E-F).

**Figure 3.**
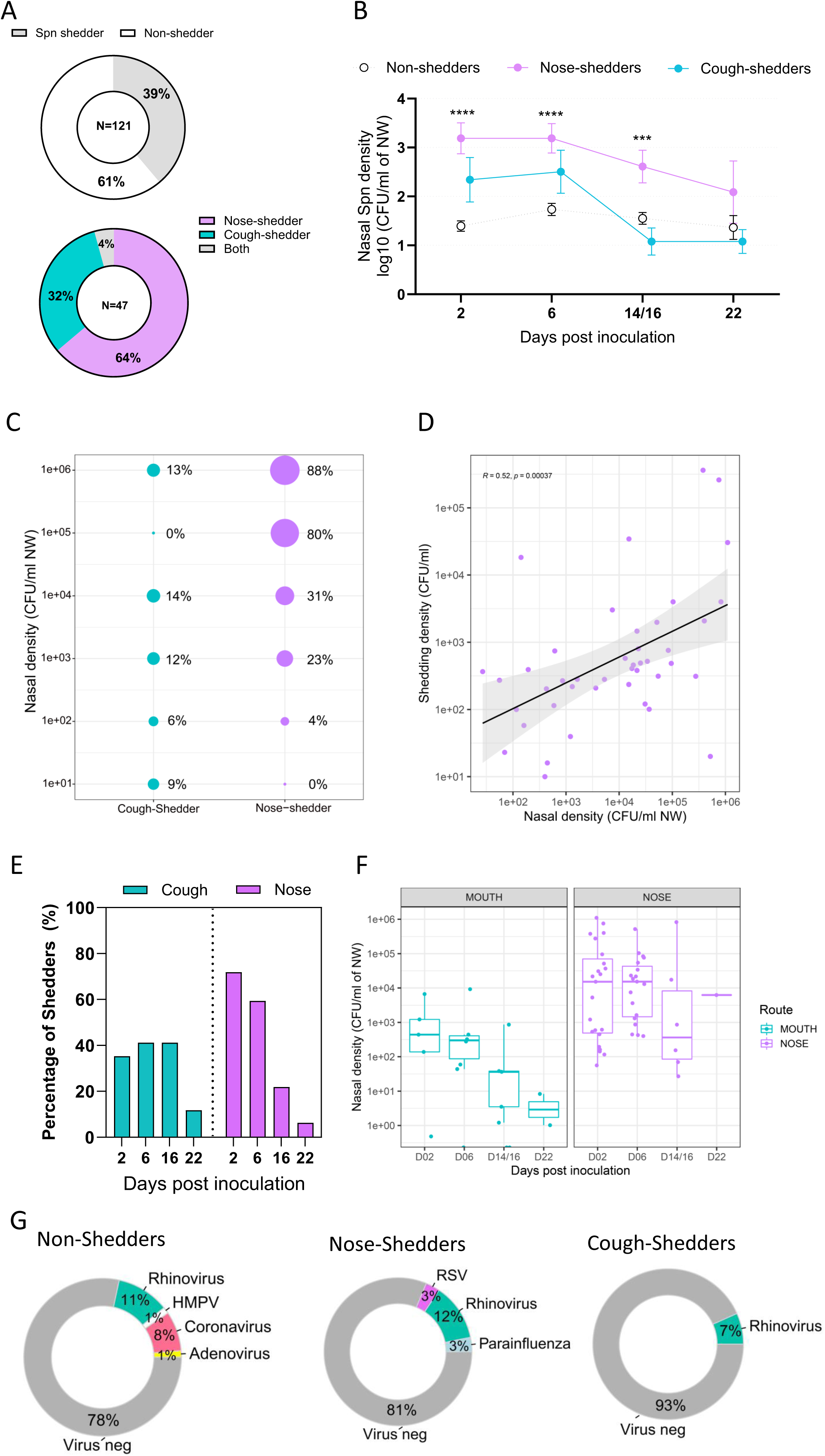
Route of bacterial shedding and association with colonising density. **A)** Percentage of bacterial shedders (n=47) and non-shedders (n=74) amongst the Spn colonised individuals (top) and breakdown for route of shedding in the Spn shedders. **B)** Mean ± SEM of nasal pneumococcal density per time point after inoculation day (D0) in Spn colonised individuals who shed bacteria through the nose (nose-shedders, n=30) or mouth while coughing (cough-shedders, n=15) or did not shed bacterial (non-shedders, n=74). **C)** Incidence of bacterial shedding for cough- and nose-shedders in association with nasal pneumococcal density expressed as log10 increased increments. **D)** Correlation between nasal Spn density (CFU/ml of NW) and shedding density (CFU/ml), Linear regression line with 95% confidence interval (grey shading) are shown. **E)** Percentage of Spn- shedders who emitted bacterial via coughing or nose per each time point after spn challenge. **F)** Levels of nasal pneumococcal density measured per time point PI in cough-shedders (turquoise) and nose-shedders (purple). Data are presented as median values and interquartile ranges (IQRs). **G)** Respiratory viral infection incidence in non-shedders, nose- and cough-shedders. Statistical differences were determined by Kruskal–Wallis test following correction for multiple comparisons (B), Fisher-test exact test (C) or two-tailed Spearman rank test (D). ***p < 0.001, ****p < 0.0001.

**Table 3:**
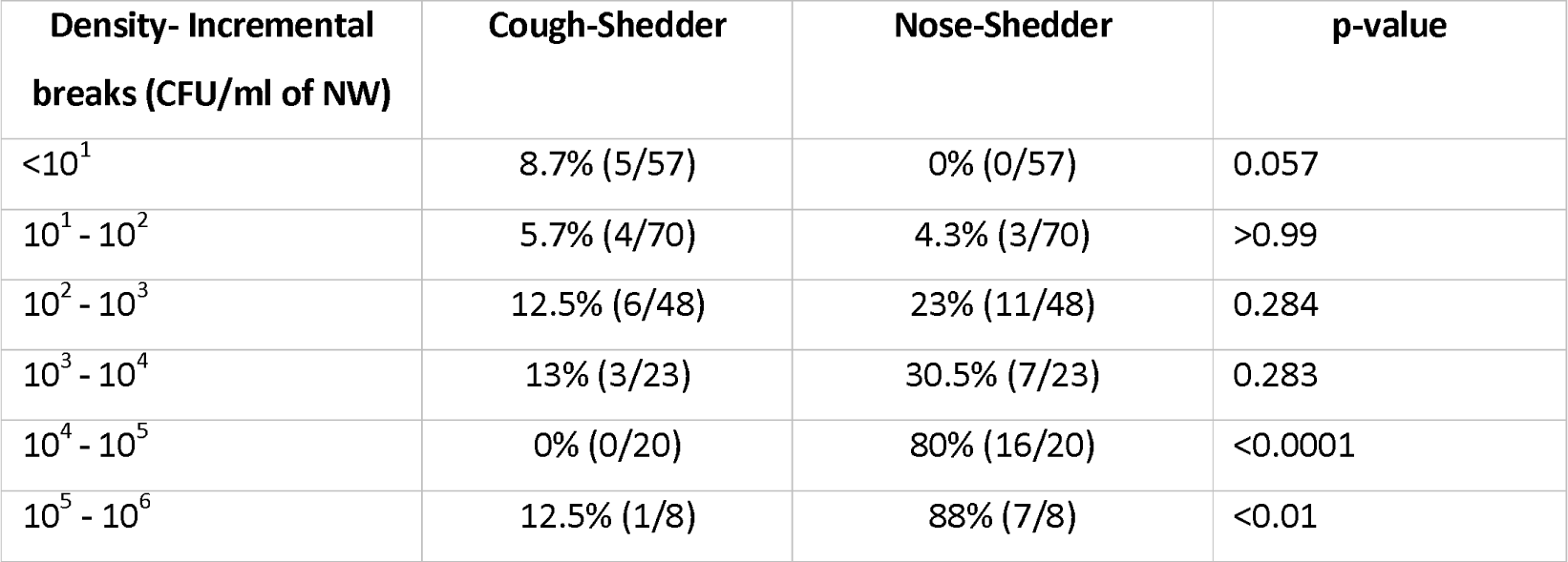
Percentage of cough- and nose-shedders per density increment.

| Density- Incremental breaks (CFU/ml of NW) | Cough-Shedder | Nose-Shedder | p-value |
| --- | --- | --- | --- |
| $<10^1$ | 8.7% (5/57) | 0% (0/57) | 0.057 |
| $10^1 - 10^2$ | 5.7% (4/70) | 4.3% (3/70) | >0.99 |
| $10^2 - 10^3$ | 12.5% (6/48) | 23% (11/48) | 0.284 |
| $10^3 - 10^4$ | 13% (3/23) | 30.5% (7/23) | 0.283 |
| $10^4 - 10^5$ | 0% (0/20) | 80% (16/20) | <0.0001 |
| $10^5 - 10^6$ | 12.5% (1/8) | 88% (7/8) | <0.01 |

We also investigated the contribution of respiratory viruses in bacterial shedding, due to previously observed associations with density changes and induction of nasal inflammatory responses. Coronaviruses was detected only in the non-shedders group in a proportion of 8% (6/74), rhinovirus was found in similar proportion in shedders and non-shedders, whereas RSV and HPIVs was detected in a small proportion (3%, 1/30) of nose-shedders only. There was no difference in viral copies detected in Spn-shedders and non-shedders (Suppl. Figure 4A). However, Spn-shedders with asymptomatic URTI had substantially higher bacterial colonising density at D2, D6 and D16 post inoculation compared to non-shedders with (Suppl. Figure 4B). The small numbers of individuals with confirmed asymptomatic URT and shedding data limited further investigation of the role of viral infection in shedding.

### Distinct nasal cell profile and gene expression patterns in nose-shedders

We next analysed the cellular composition of nasal cells obtained by nasal microbiopsies, applying flow cytometry. The number of leukocytes, granulocytes and monocytes was expressed as a ratio to epithelial cells [20] and compared between the three Spn-colonised groups at baseline, day2, day6 and day16 post pneumococcal challenge (Figure 4A). At baseline, overall levels of nasal immune cells were comparable amongst the groups, with the exception of CD8^+^ T cells levels (Figure 4A). After challenge, nose-shedders had a substantial increase in the proportion of neutrophils compared to non-shedders and in monocytes and CD4^+^ T cells compared to both cough- and non-shedders. This increase in immune cells was observed only at 2 days post challenge and only in the nose-shedders group.

**Figure 4.**
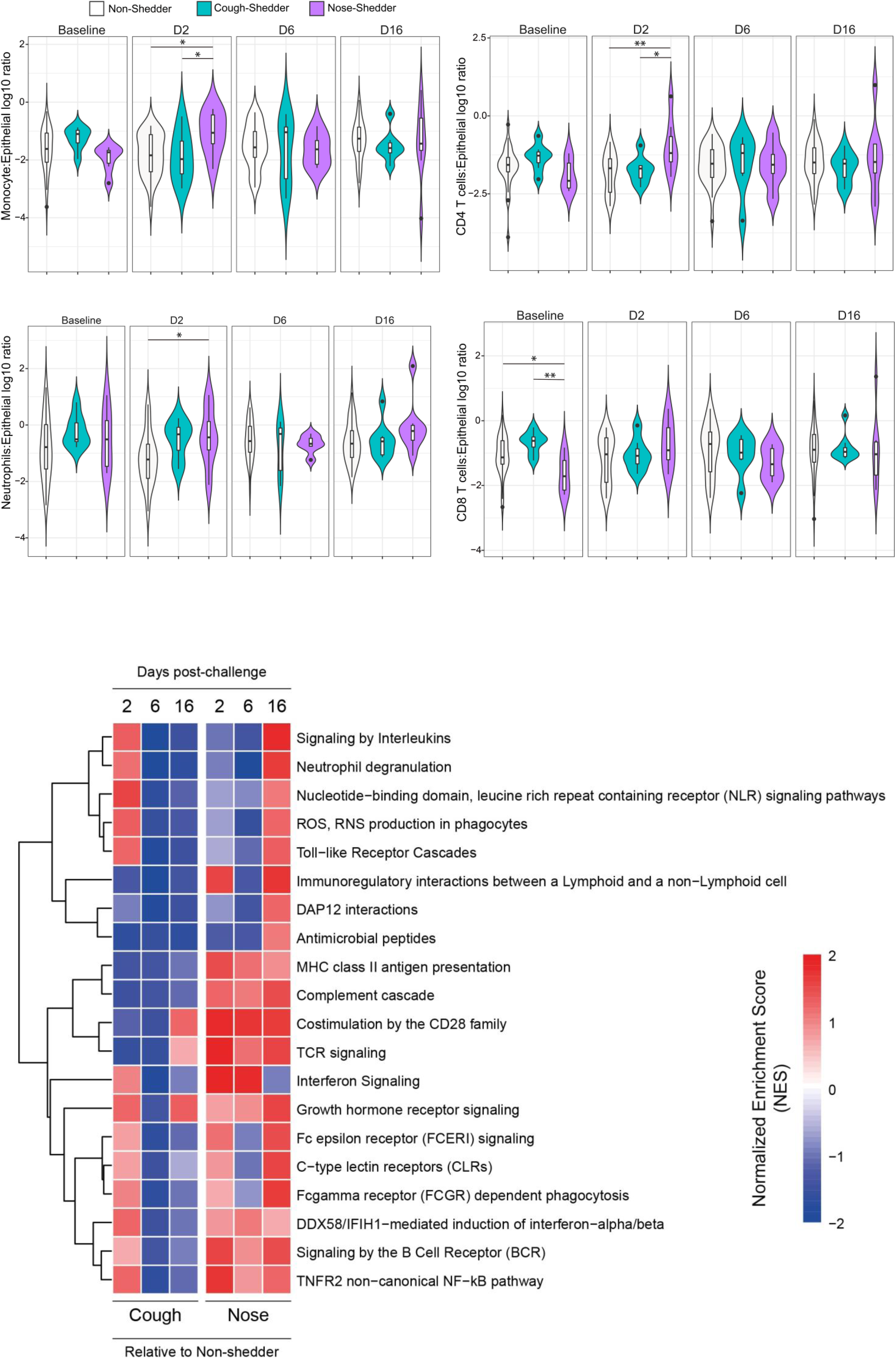
Distinct nasal inflammatory profile in those with asymptomatic URT infection. **A)** Volcano plots showing the median and IQR of nasal monocyte, neutrophils and T cell numbers normalised to the epithelial cell numbers in non-shedders (n=), cough-shedders (n=) and nose-shedders (n=). Statistical differences were determined by Kruskal–Wallis test following correction for multiple comparisons. B) Gene set variance analysis (GSVA) of immune response pathways of nasal cells collected at D2, D6 and D16 after pneumococcal in cough- and nose-shedders in relation to non-shedders colonised individuals at the same time points. Normalized enrichment score (NES) is presented in gradient colour. Red shades indicate pathways overrepresented, whereas blue shades depict the underrepresented pathways at each time point in relation to non-sheders group. Statistical differences were determined by Wilcoxon’s paired test corrected by multiple-comparison testing (Benjamini-Hochberg).

Nasal cells were also obtained from the same individuals for a parallel transcriptome analysis (Figure 4B). Genes transcriptional changes in cough- and nose-shedders per time point post challenge were described in relation to non-shedders as the reference group. Cough-shedders had an enrichment in gene sets associated with innate immune responses and cytokine signalling at the very early stages of pneumococcal colonisation (day 2 PI), which was dismissed at later time points. In nose-shedders, adaptive immunity gene sets and cytokines signalling were upregulated early after colonisation, whereas enrichment in innate immune system gene sets was delayed (day16 PI).

### Nasal inflammation drives bacterial shedding through the nose

Nasal inflammation was also assessed by measuring the levels of 30 cytokines in the nasal fluid of nose- and cough-shedders at four time points (D2, D6, D16 and D22) post-challenge and compared with the levels measured in non-shedders at the same time points. Nose-shedders had a distinct pro-inflammatory profile, defined by enrichment in growth factors, pro-inflammatory cytokines (IL-1b and TNFa) and chemokines that affect monocytes and granulocytes migration, such as macrophage-inflammatory protein-1a and -1b (MIP-1a, MIP-1b) and granulocyte-colony stimulation factor (G-CSF). This increase in nasal inflammatory responses compared to non-shedders group was observed at D2 (upregulation of 9 out of 30 cytokines), D6 and D16 (upregulation of 7 out of 30 cytokines) but not at D22 post-challenge. On the other hand, cough-shedders did not differ from their non-shedder counterparts at any time point over the course of experimentally induced pneumococcal colonisation.

Acute nasal inflammation was identified as a key characteristic of nasal bacterial shedding, as in the case of pneumococcal density. To dissect the associations between nasal inflammatory responses, bacterial colonising density and bacterial shedding, these parameters were analysed using correlation plots for the time points wherein bacterial shedding was more frequently observed (D2 and D6 post-infection). Notably, correlation plots indicated a positive association between nasal inflammatory responses and bacterial shedding through the nose. At day 2 post-challenge, the association with inflammatory responses included pro-inflammatory cytokines (IL-6, TNFa, IL-1RA) and the involvement of several monocyte and granulocytes chemokines and growth factors [monocyte-chemoattractant protein-1 (MCP-1), MIP-1a, MIP-1b, MIG and granulocyte-monocyte colony stimulation factor (GM-CSF)] (Figure 5C), whereas at day 6 associations were restricted to only 2 pro-inflammatory cytokines (IL-6 and TNFa) (Figure 5D). A similar relationship was also observed between nasal inflammation and pneumococcal density, deriving mainly from pro-inflammatory responses and MCP-1 chemokine at Day 2 (Figure 5C) and was enriched with additional chemoattract cytokines (MIP-1a, MIP-1b, RANTES) at Day 6 post-challenge (Figure 5D).

**Figure 5.**
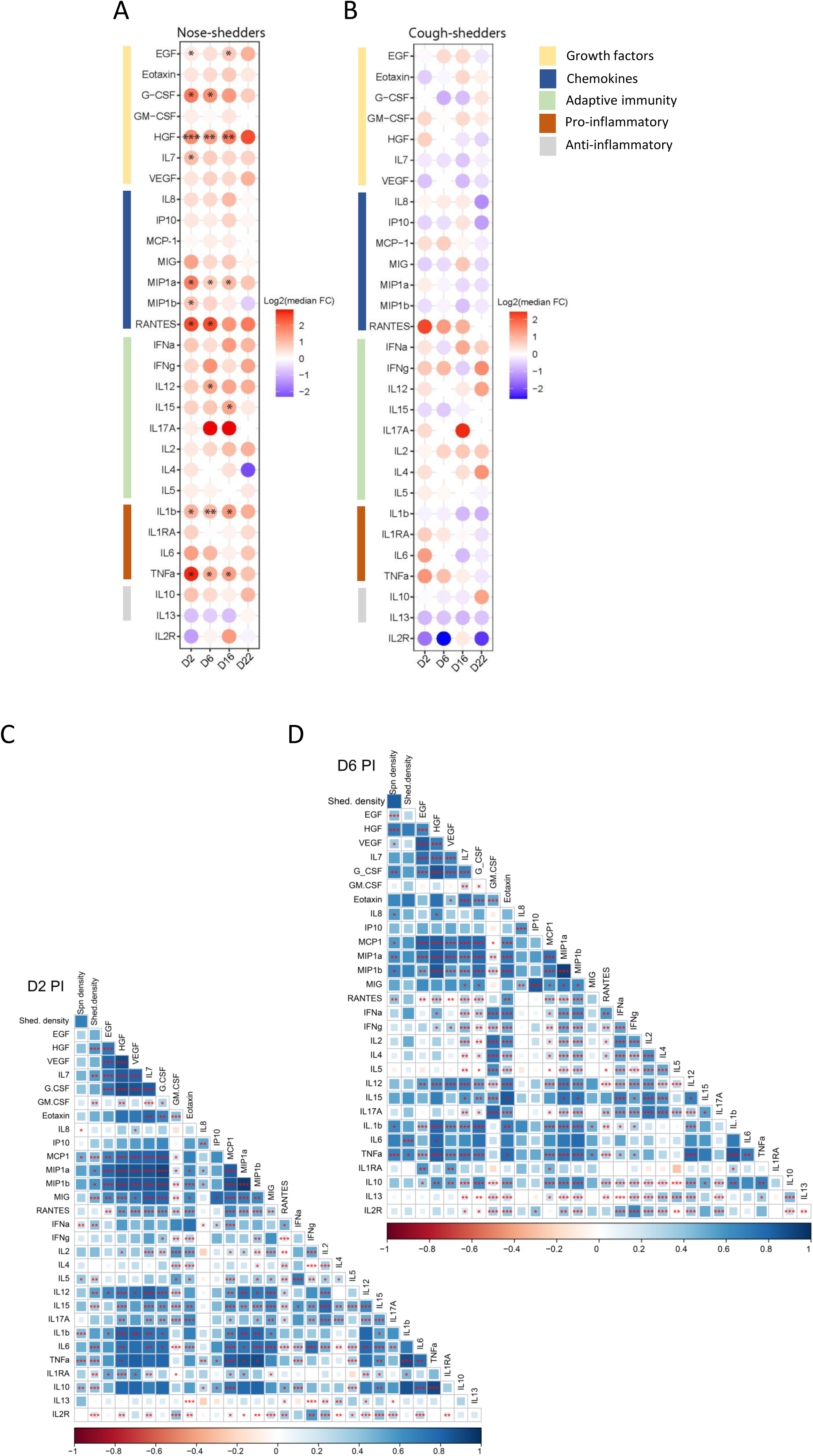
Nasal inflammation in nose-shedders and correlation bacterial density and shedding. Heat maps showing the median log_2_ fold change (FC) in the levels of 29 cytokines and chemokines in the nasal wash samples **a)** nose-shedders (n=28) and **b)** cough-shedders (n=8) compared to non-viral infected group (n = 46) at D2, D6, D16 and D22 post challenge. Used two-tailed Mann-Whitey test, followed by Benjamini-Hochberg correction for multiple testing**. C-D)** Correlograms of nose-shedders at D2 and D6 post-challenge, respectively. Spearman rank order correlation values (R) are shown from red (−1.0) to blue (1.0); R values are indicated by colour and square size. P values are indicated by red asterisks. *p < 0.05, **p < 0.01, ***p < 0.001.

## DISCUSSION

In humans, respiratory viral and pneumococcal co-infections have mainly been studied in the context of disease (hospitalisation cases, pathogenicity) [10, 38] describing associations rather than a causative relationship. How respiratory viruses influence pneumococcal carriage acquisition, leading to subsequent disease and bacterial transmission to the community, is not well described.

Here, we investigated the impact of natural viral URTI on acquisition of experimentally induced colonisation and have shown that primary infection with certain respiratory viruses, such as rhinovirus, RSV, PIVs, but not seasonal coronaviruses, increased pneumococcal carriage rates. In agreement with previous reports[39, 40], presence of these respiratory viruses in the upper airways also led to higher pneumococcal colonising density, with RSV infection having the most striking effect. It is possible that viral infection and subsequent inflammation provide an environment, enriched in nutrients, promoting bacterial proliferation [41].

Our results support epidemiological and clinical evidence, suggesting that certain respiratory viruses, may have a substantial role in the burden of pneumococcal disease [8, 9, 38, 42–44]. A newly acquired carriage episode poses an increased risk towards disease development, likely because the host has not yet developed immunity to the pathogen[45, 46]. Hence, a viral-induced pneumococcal colonisation episode, which commences with high colonisation density, could trigger disease development, particularly in the at-risk populations, such as the very young, the old and the immunocompromised.

RSV significant contribution to paediatric IPDs was recently highlighted by two surveillance studies contacted in Israel[8] and France[9] during implementation of non-pharmaceutical interventions early in COVID-19 pandemic. Both studies reported that the overall prevalence of pneumococcal carriage in paediatrics was not significantly altered. This could be a result of population recruitment, study set-up and time of pneumococcal infection. Our study included healthy non-colonised adults, who were experimentally exposed to pneumococcus at a known time, whereas the Israel and French studies were both pneumococcal colonisation and IPD surveillance studies in children. RSV also has been linked with increased pneumococcal invasiveness. Epidemiological data have reported increased frequency of less invasive, non-vaccine included, pneumococcal serotypes (15B/C, 33F), during RSV infection in paediatric cases of community-acquired alveolar pneumonia [47]

On the other hand, there are conflicting data on the relationship between rhinovirus and parainfluenza infection and pneumococcal disease[8, 38, 39]. The balance swifts depending on the reported population, as those viral infections commonly associate with pneumococcal disease in asthmatics, HIV infected individuals and the elderly[38]. However, rhinoviral infection is very common amongst children and causes rhinitis[48], which in turn can lead to increased nasal inflammation, and hence pneumococcal shedding and onwards transmission during co-infection.

We also examined bacterial shedding through the nasal and oral route during experimentally induced pneumococcal carriage. We have showed that pneumococci exit the host more frequently via the nose, early after establishment of colonisation, predominantly when the nasopharynx is inflamed and colonised with high bacterial load (Figure 6). In animal models, modulation of nasal inflammation alters pneumococcal shedding and transmission rates, indicating a direct relationship of positive association[19, 49]. Pneumolysin, a toxin produced and secreted by pneumococci, has also been reported as a significant factor that promotes inflammation and subsequently increased bacterial shedding[49]. Our results suggest that nasal shedding is more likely to occur during the early stages of a colonisation episode, when host innate immune responses, such as local inflammation, neutrophils degranulation and phagocytes recruitment are at their peaks, as previously observed in the human challenge model[20, 50]. An inflamed nasal mucosa can lead to increased volume and flow of secretions locally, promoting shedding and prolonged pneumococcal survival in the environment [6, 49]. On the other hand, cough-induced shedding was dissociated from host responses and bacterial density (Figure 6), allows to speculate that this route of shedding is more common in older adults, wherein pneumococci can be more frequently detected in the throat and oral cavity. In this study any association between viral URLT and bacterial shedding was prohibited by the low numbers of viral-infected individuals in the nested shedding study, although rhinoviral URLT was well represented in the shedders group. Also, viral-infected shedders were colonised with higher pneumococcal density compared to their non-shedder counterparts. Our findings suggest that pneumococcal colonised adults, even when asymptomatic, may be a source of contagion, particularly when are heavily colonised, as in the case of viral co-infection.

**Figure 6.**
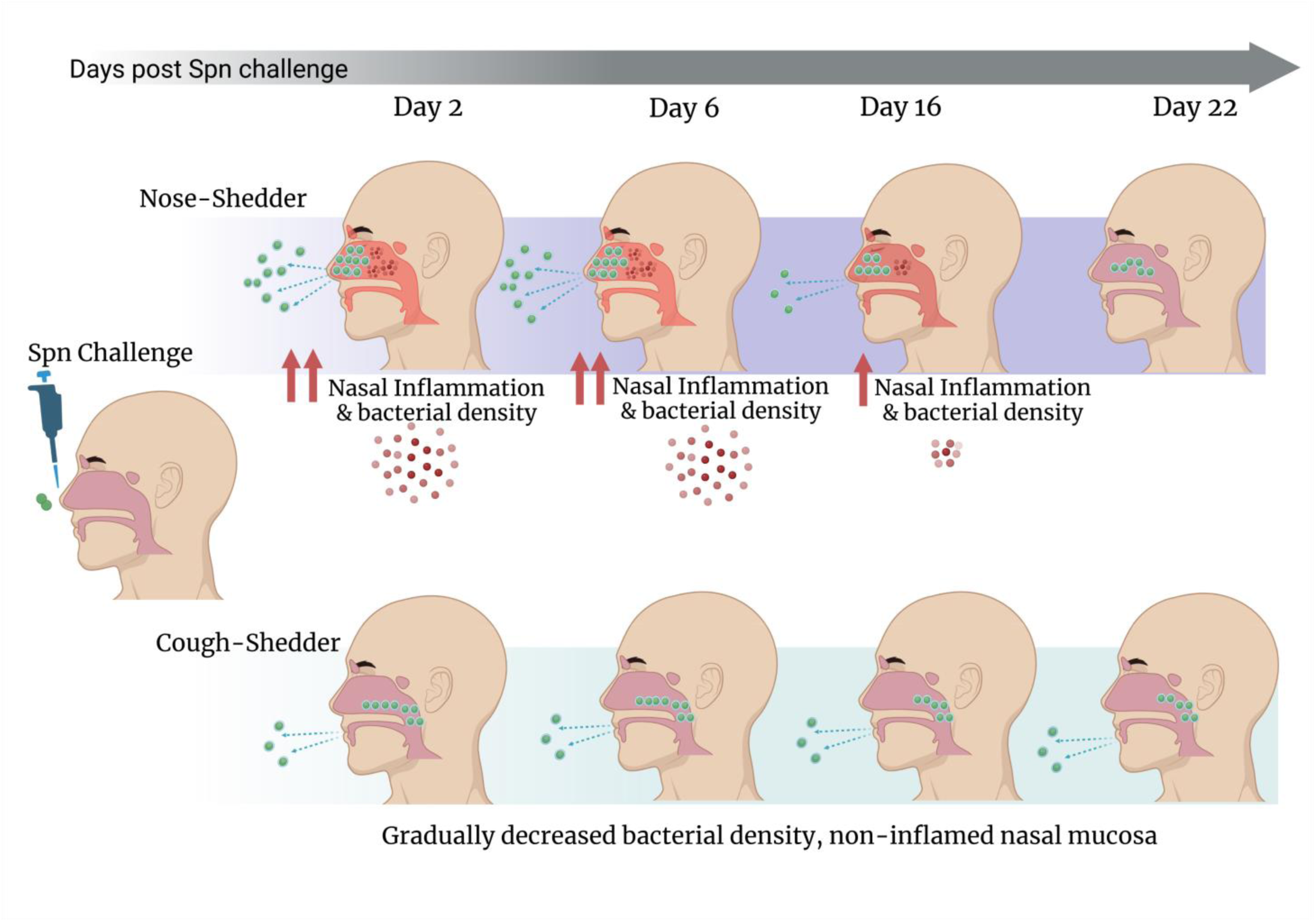

Our study has limitations. Viral URTIs were naturally acquired and assessed by molecular methods only before challenge, and hence co-infection with pneumococcus may had occurred at any time point through the course of the primary viral infection. In addition, only individuals with asymptomatic URTI were enrolled into the challenge studies, as per studies inclusion/exclusion criteria. The non-specified time of viral infection onset and the lack of symptoms may have masked viral-induced nasal inflammation and the optimum effect that certain respiratory viruses have on carriage acquisition. However, our results reflect some of the viral-pneumococcal associations described in large epidemiological studies, as in the case of highly influential RSV[8, 10, 51, 52] and the ambiguous contribution of seasonal coronaviruses in secondary pneumococcal diseases [53, 54]. Another limitation arises from the monomeric use of only pneumococcal serotype 6B, as challenge agent. Further studies are needed to investigate whether viral-pneumococcal relationship is serotype dependent and whether during colonisation with other pneumococcal serotypes, the bacterial exhibit the similar or differential pattern of shedding. Another limitation is that we only investigated one direction of viral-bacterial relationship, with virus infection coming first. Further studies are needed to investigate the effects of a concurrent virus infection in an established colonisation episode as well as how pneumococcus can impact on subsequent virus infections and host responses.

The findings of this study suggest that protection against respiratory viral infections and control of pneumococcal density would have a major contribution in preventing pneumococcal disease and blocking bacterial transmission. Vaccine interventions have the potential to confer this dual outcome. Current licenced pneumococcal conjugate vaccines have undoubtedly reduced IPD cases[55], but their effect on pneumococcal colonisation density has been underestimated. Control of pneumococcal density by PCVs, as shown in experimentally colonised adults[56], can play a pivotal role in preventing pneumococcal disease and reducing pneumococcal transmission. Yet, PCVs protect against a limited number of the most invasive serotypes. After interaction with certain viruses, less invasive, non-vaccine serotypes may exhibit heightened ability to invade or exit the human host. In this occasion, viral vaccines, may further contribute to the reduction of pneumococcal disease burden and onwards transmission as an additional benefit beyond the virus-specific direct protection.

## Data Availability

All data produced in the present study are available upon reasonable request to the authors

## Acknowledgments

We thank all the patients and healthy volunteers who participated in the present study and all the clinical staff who helped with recruitment and sample collection. The study was financially supported by the Bill and Melinda Gates Foundation (grant no. OPP1117728) and the UK Medical Research Council (grant no. M011569/1) awarded to D.M.F and S.B.G. and a Robert Austrian Research award awarded to E.M.

## Authors Contribution

E.M and D.M.F conceived and designed the study. H.H, A.H.W., M.F., R.R and A.M.C. recruited, consented study participants and obtained human samples. E.N, A.B, A.H, J.R, F.E and C.S processed samples. E.M, E.N., A.G, J.R and D.B generated and analysed the data.

E.M., A.G. and D.M.F interpreted data. E.M. wrote the manuscript and with subsequent inputs from the co-authors. All co-authors approved the final version of the manuscript.

