## Supplementary material for "RSV and Rhinovirus asymptomatic upper airway infection increases pneumococcal carriage acquisition rates and density in adults whereas nasal inflammation is associated with bacterial shedding"

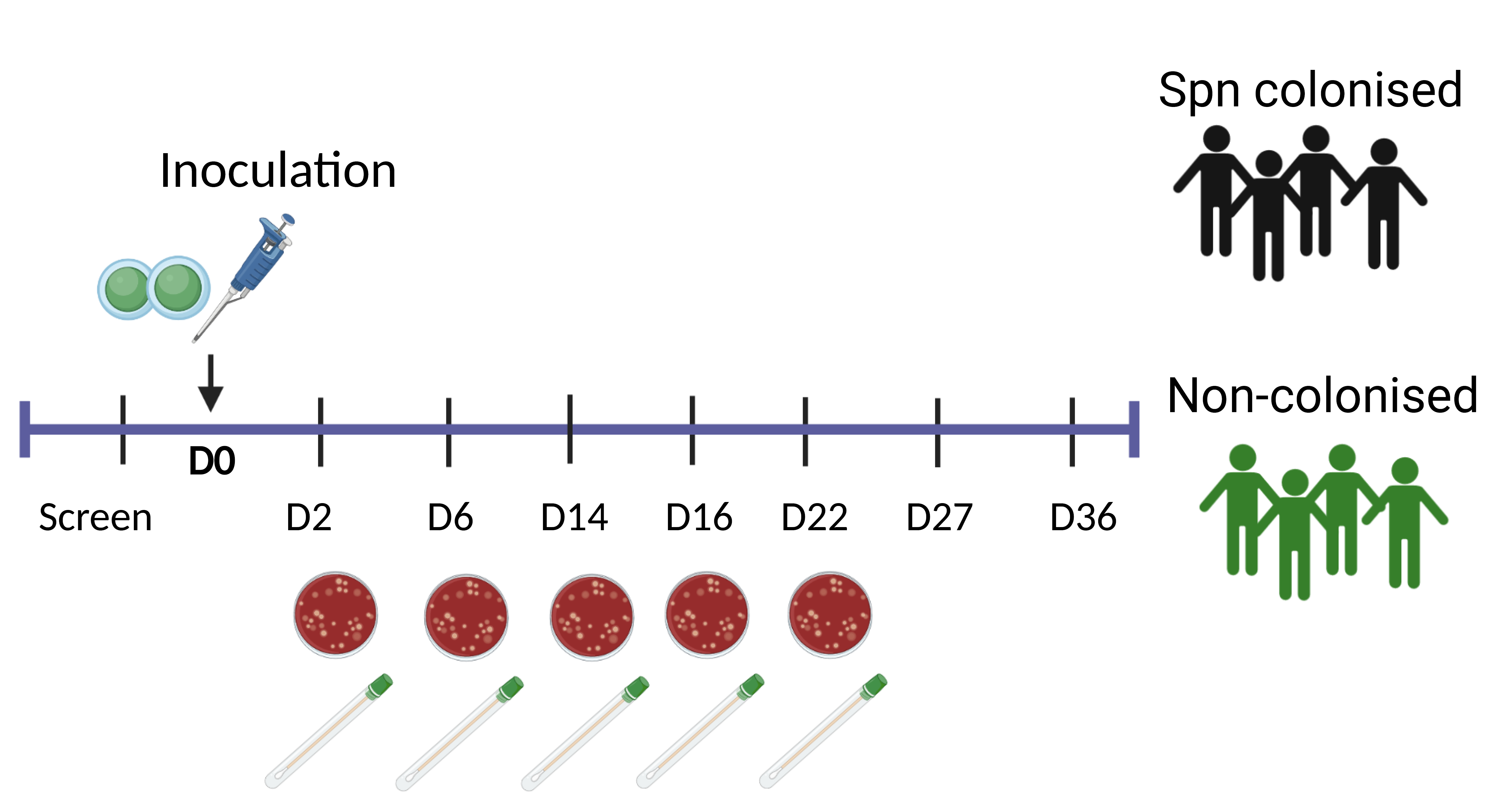

**Supplementary figure 1: Study design for assessing bacterial shedding in pneumococcal challenged adults.** Collection of cough plates and nose-to-hand swabs at several time points post pneumococcal challenge for both Spn-colonised (n=121) and non-colonised subjects (n=48).

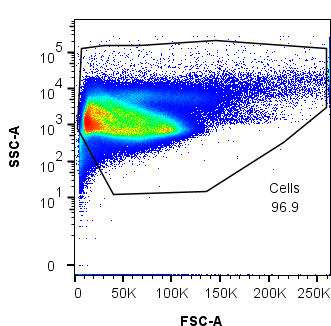

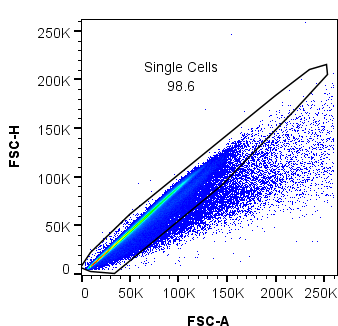

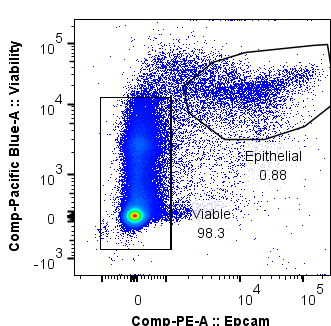

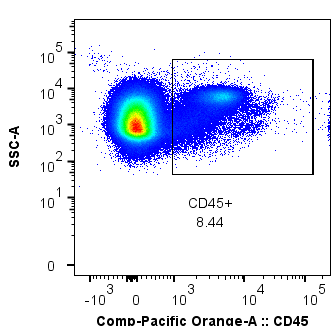

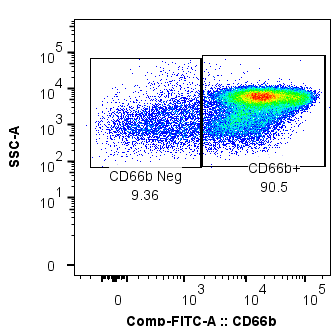

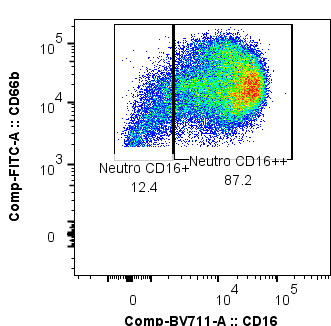

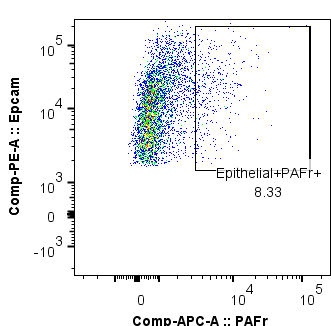

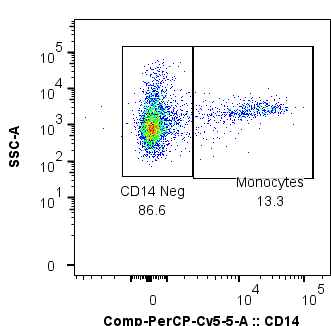

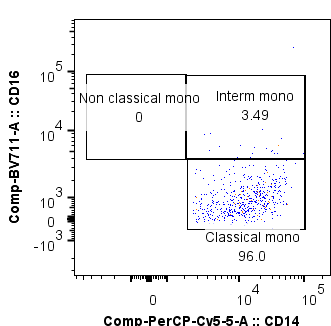

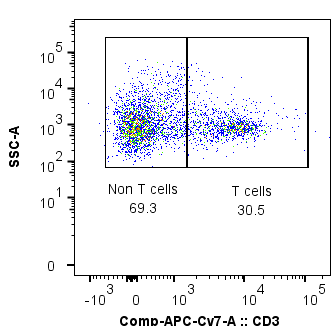

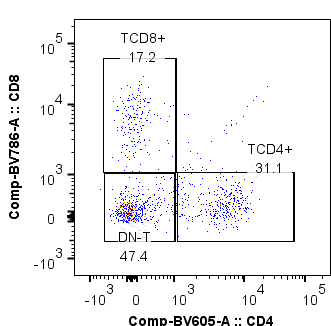

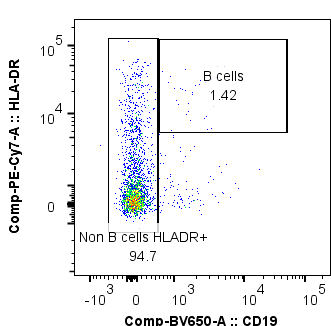

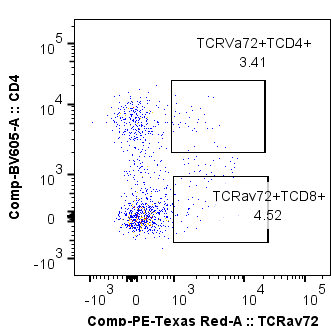

**Immune cells**

**Epithelial cells**

**Supplementary Figure 2. Gating strategy from a representative nasal cell sample by flow cytometry. (A)** Identification of the major nasal cellular subsets obtained from nasal curettage by flow cytometry: after excluding debris and doublets, the epithelial cells were identified by Epcam expression. A viability dye and CD45 were used to identify live immune cells. Among those cells, side scatter, CD66b, CD14, CD19 and CD3 were used to identify nasal granulocytes, monocytes, B and T cells respectively. Subsequently, we also analysed different subsets within each immune cell population for a further characterisation of the nasal cell populations.

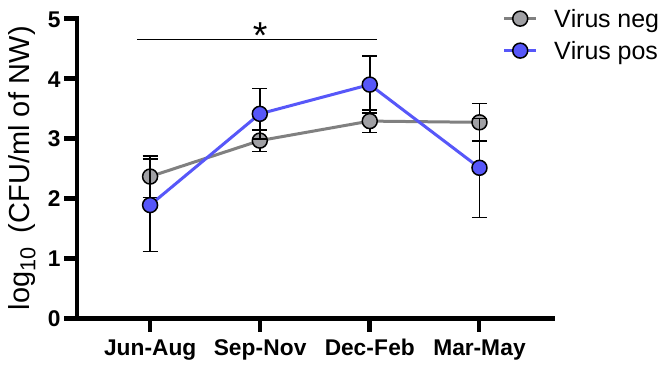
**A	B**

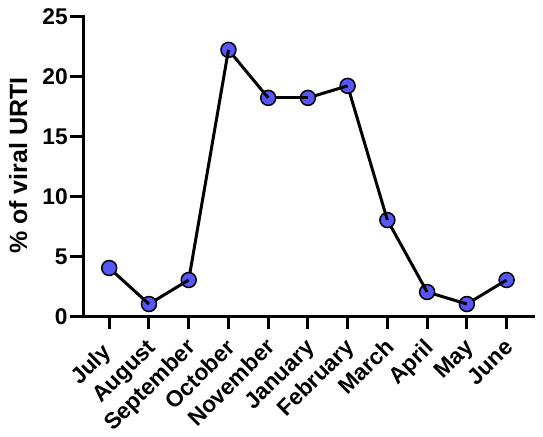

**Supplementary figure 3: Seasonality effect on incidence of URTIs and pneumococcal colonising density. A)** Frequency of asymptomatic upper airway viral infection per calendar month (n=99 virus infected individuals). **B)** Mean ± SEM of pneumococcal density per season in virus co-infected pneumococcal carriers (blue) and non-infected pneumococcal carriers (in grey). *p < 0.05

**A	B**

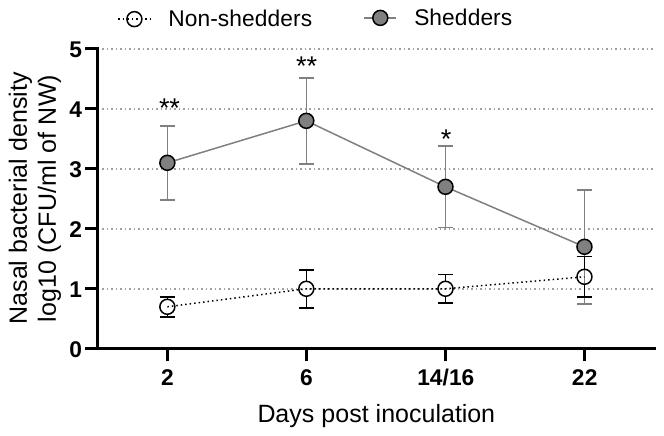

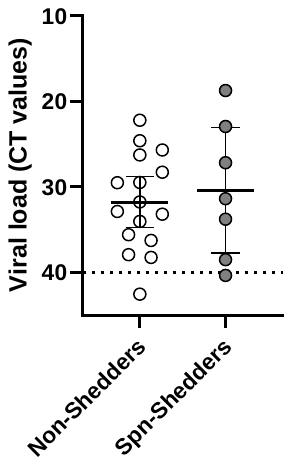

**Supplementary figure 4: Bacterial shedding and respiratory viral infection. A)** Levels of detectable viral load, expressed as CT values, in nasopharyngeal samples from non-shedders (n=16) and Spn-shedders (n=7). **B)** Mean ± SEM of nasal pneumococcal density in non-shedders (n=16) and Spn-shedders (n=7) with detectable viral-infection. Densities were calculated from classical microbiology (log10[cfu/ml +1]). Statistical differences were determined using Welch t-test (B) and ANOVA test (C). *p < 0.05, **p < 0.01.

| **Marker** | **Fluorochrome** | **Clone** | **Provider** | **Reference** |
| --- | --- | --- | --- | --- |
| TCRVα7.2 | PE/Dazzle594 | 3C10 | Biolegend | 351730 |
| CD14 | PerCP.Cy5.5 | MφP9 | BD Biosciences | 562692 |
| CD16 | BV711 | 3G8 | Biolegend | 302044 |
| CD19 | BV650 | HIB19 | Biolegend | 302238 |
| CD3 | APC-Cy7 | SK7 | Biolegend | 344818 |
| CD45 | BV510 | HI30 | Biolegend | 304036 |
| CD4 | BV605 | SK3 | Biolegend | 344646 |
| HLADR | PECy7 | L243 | Biolegend | 307616 |
| EpCAM | PE | 9C4 | Biolegend | 324206 |
| CD8 | BV785 | SK1 | Biolegend | 344740 |
| CD66b | FITC | G10F5 | Biolegend | 305104 |
| PAFr* | APC | 11A4 -21/NA | Cayman/Abcam | 160600/ab201807 |
| Live&Dead | PB | NA | ThermoFisher | L34964 |

**Supplemental table 1. Summary and specifications of the multiparametric flow cytometry antibodies and viability dye used for nasal cells immunophenotyping.** We carefully developed a multiparametric flow cytometry panel composed with twelve different monoclonal antibodies (from Biolegend, Cayman Chemical and BD Biosciences) including a viability dye (ThermoFisher). PAF receptor purified monoclonal antibody was conjugated with APC using an APC-Conjugation kit (Abcam). Electronic compensation was set using CompBeads (BD Biosciences), Arc beads (ThermoFisher according to manufacturer’s instructions), and samples were acquired into a BD LSR2 cytometer (BD Biosciences). Abbreviations (NA, Not applicable).
